# A Status-Neutral Approach to HIV – Is Targeted Testing Still Relevant South of Sahara?

**DOI:** 10.1101/2024.04.16.24305893

**Authors:** Hamufare Mugauri, Owen Mugurungi, Joconiah Chirenda, Kudakwashe Takarinda, Prosper Mangwiro, Mufuta Tshimanga

## Abstract

**Introduction:** In 2022, UNAIDS replaced the 90% Global HIV targets with six Comprehensive 95% targets that include linkage to comprehensive HIV prevention services, the thrust of the status-neutral approach to HIV testing. Zimbabwe has been implementing both targeted testing and the status-neutral concept. In this paper, we analyse the role of status-neutral concepts in targeted testing, for effective case identification and linkage to prevention and treatment services.

**Methods:** We conducted a cross-sectional study on 36 multi-stage sampled sites across 4/10 provinces of Zimbabwe. Screened and non-screened patients were tested and analysed for positivity ratios and linkage to post-test services. Data were extracted using Epicollect5 and imported into EpiData software and Stata for cleaning and analysis. Data were summarized as proportions, odds ratios and adjusted odds ratios at 5% significance level.

**Results:** Of 23,058 HIV tests done, females constituted 55% (n=12,698), whilst 63.5% (n=14,650) were retests and positivity of 7.5% obtained. Screened patients contributed 75.1% to the overall positivity (1,296/1,727), from 66% (n=15,289) of the total tests conducted. The 45–49-year category was 3.6 times more likely to test positive (a95%CI:2.67,4.90). Males were 3.09 times more likely to test positive in adjusted analysis (a95%CI: 2.74, 3.49), from an 8% (n=912) positivity ratio. First tests were 65% more likely to test HIV positive (a95%CI: 1.43, 1.91) whilst screened patients were 3.89 times more likely to link to HIV prevention services (a95%CI: 3.05, 4.97), against 25.5% (n=1,871) linkage among patients not screened

**Conclusion:** Targeted and status-neutral testing are related and complimentary concepts which, when simultaneously applied, potentiates case identification through prioritizing high-risk individuals for testing, as well as arresting ongoing transmission of HIV through effective linkage to HIV prevention and treatment. This approach facilitates economic usage of limited resources, in generalized epidemics.

## Introduction

Status-neutral HIV testing is a novel approach to HIV education, testing and treatment that accentuates a continuum of care regardless of the individual’s HIV diagnosis [1]. The concept infers that all people, regardless of their HIV status, are treated in the same way from the start of the HIV testing process and linked to appropriate services based on their test results. Further, it envisages improved health outcomes, the prevention of new infections and the establishment of a world where HIV is untransmissible through prevention and treatment options availed to everyone [2].

The concept also focuses on activities that meet the needs of populations at risk or living with HIV, rather than dividing services into prevention or care. Status-neutral approach to HIV prevention and care defines the entry point to care as the time of an HIV test [3]. At this point, clients’ needs are assessed, and they are engaged and linked to appropriate services based on these needs, regardless of whether their HIV test is positive or negative [4].

First introduced by the New York City Department of Health and Mental Hygiene in 2016, this new paradigm is a comprehensive system of prevention that includes all people affected by HIV, regardless of their HIV status [5–7]. It further highlights elaborate steps that can lead to an undetectable viral load and steps for effective combination HIV prevention. The concept is also premised on the assertion that most countries have achieved the 95% targets and therefore should be focused on consolidating these achievements by giving equal attention to negative testers, who were largely neglected in the quest of chasing positivity ratio targets [8].

The concept is rapidly being embraced globally with the World Health Organization (WHO) and Centers for Disease Control promoting it through various for [9]. Many countries have since adopted the concept of variability influenced by context and needs.

Zimbabwe, located in sub–Saharan Africa, is one of the countries severely affected by the HIV pandemic, where HIV remains firmly established as a generalized epidemic with a prevalence of 11,01% and incidence of 17% translating to approximately 23,000 new infections every year [10, 11]. (**Figure 1**)

**Figure 1:**
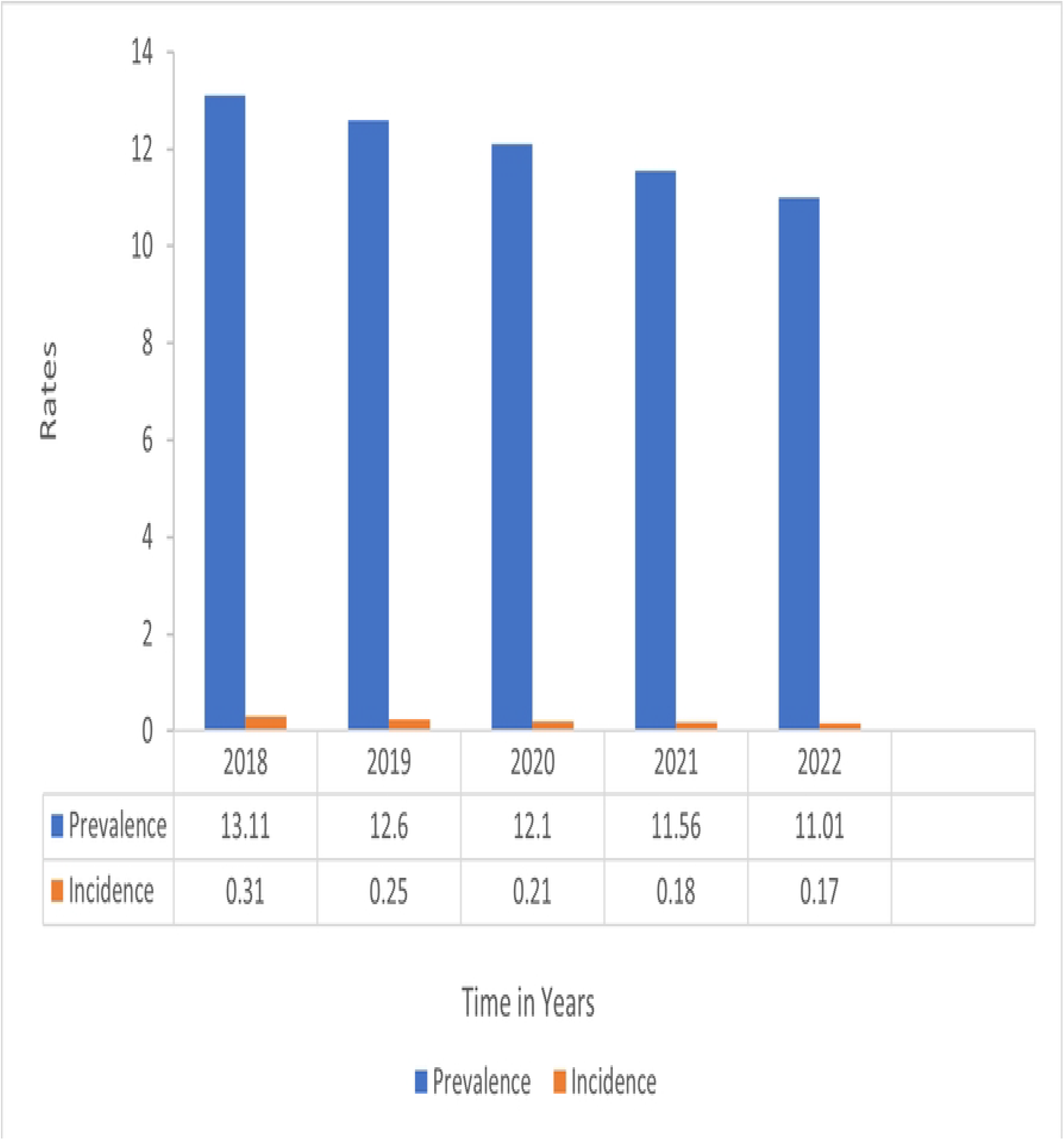
HIV Prevalence and Incidence Rates in Zimbabwe, 2018-2022 (Source: UNAIDS HIV Estimates)

To address her predicament, Zimbabwe has been implementing a Targeted testing model that prioritizes high-risk individuals through a screening algorithm, adopting differentiated testing models that include strengthening Index testing-a proven high-yield HIV testing model [12].

Whilst, remarkably, the country has achieved the 95% targets, according to UNAIDS HIV estimates, the country still bears a generalized epidemic contributed by population and geographically varied sub-epidemics requiring innovations for case finding and effective HIV prevention packages [13].

This context demands that Zimbabwe, like her Southern African counterparts, needs interventions designed to arrest ongoing transmission of HIV, identify the remaining cases, and put them on effective life-long treatment to bring the pandemic to an end. This paper therefore interrogates the relevance of Status neutrality in the context of a generalized epidemic, high new infections and established ongoing transmission of HIV to recommend the context-specific application of the novel status-neutral concept.

## Materials and Methods

### Study Design

We conducted a cross-sectional study, with an analytical component.

### Setting

#### General setting

Zimbabwe is a landlocked, low-income country in Southern Africa located between Botswana, South Africa, Mozambique, and Zambia with an estimated population of 16,3 million and a human development index of 0.593, ranked number 174 globally out of 189 countries in 2022[14, 15]. The country is divided into two urban provinces, eight rural provinces and 62 districts.

#### Zimbabwe National HIV Programme

The AIDS and TB Programme (ATP) is mandated to coordinate the development of HIV/AIDS health policies and set up national standards and guidelines as part of the national response to HIV in Zimbabwe. Four sub-units under ATP, namely, HIV Prevention, Care and Treatment, Prevention of Mother to Child Transmission (PMTCT) and Monitoring and Evaluation (M & E). These sub-units are delegated to ensure seamless yet specialised programming to ensure adequate response to the pandemic [16].

The HIV Prevention program oversees the activities of HIV Testing Services (HTS) activities with the ATP. Since 2016, the HTS programme has been pursuing targeted testing as an approach to reduce testing volumes, increase efficiency in HIV testing and enhance the identification of people living with HIV, to enrol them on life-long Antiretroviral Therapy (ART). Eligibility for HIV testing is done using a validated Screening algorithm [17]. In this algorithm, high-risk individuals are offered provider-delivered testing whilst those screened out are offered HIVST kits for self-screen. Following a negative test result, the patient is further screened for eligibility for combination prevention which includes PrEP.

#### Specific Study Site

The study sites were four provinces, selected out of 10 provinces in Zimbabwe. Manicaland is a province in eastern Zimbabwe. After Harare Province, it is the country’s second-most populous province, with a population of 1.75 million, as of the 2012 census [18]. Mashonaland West Province Mashonaland West is located to the North of Zimbabwe and shares the international border with Zambia in the North. Internally, the borders of the province are with Midlands Province in the West, Matabeleland North in the West, Mashonaland Central in the East, Harare, and Mashonaland East in the Southeast [19].

Matabeleland South Province covers the south-eastern plateau of Zimbabwe, and it stretches to the Botswana border on the east and borders South Africa on the South [20]. Midlands province has an area of 49,166 square kilometres and a population of 1,614,94 [21].

### Client population

All clients who were tested for HIV and documented in HIV Testing registers, at the 36 sampled facilities between 1 October 2023 to 31 December 2023 were included in the study.

### Sampling

Multi-stage sampling was done to randomly select 4 out of 10 Zimbabwe provinces using the lottery method. Further, 3 districts per province were randomly selected resulting in 12 districts. All health facilities of the 12 districts were included in the study resulting in a total of 36 health facilities being included. The sampling criteria aimed to achieve a balanced representation of health facilities, resulting in a mix of high and low volume, as well as urban and rural sites. **(Table 1)**

**Table 1.**
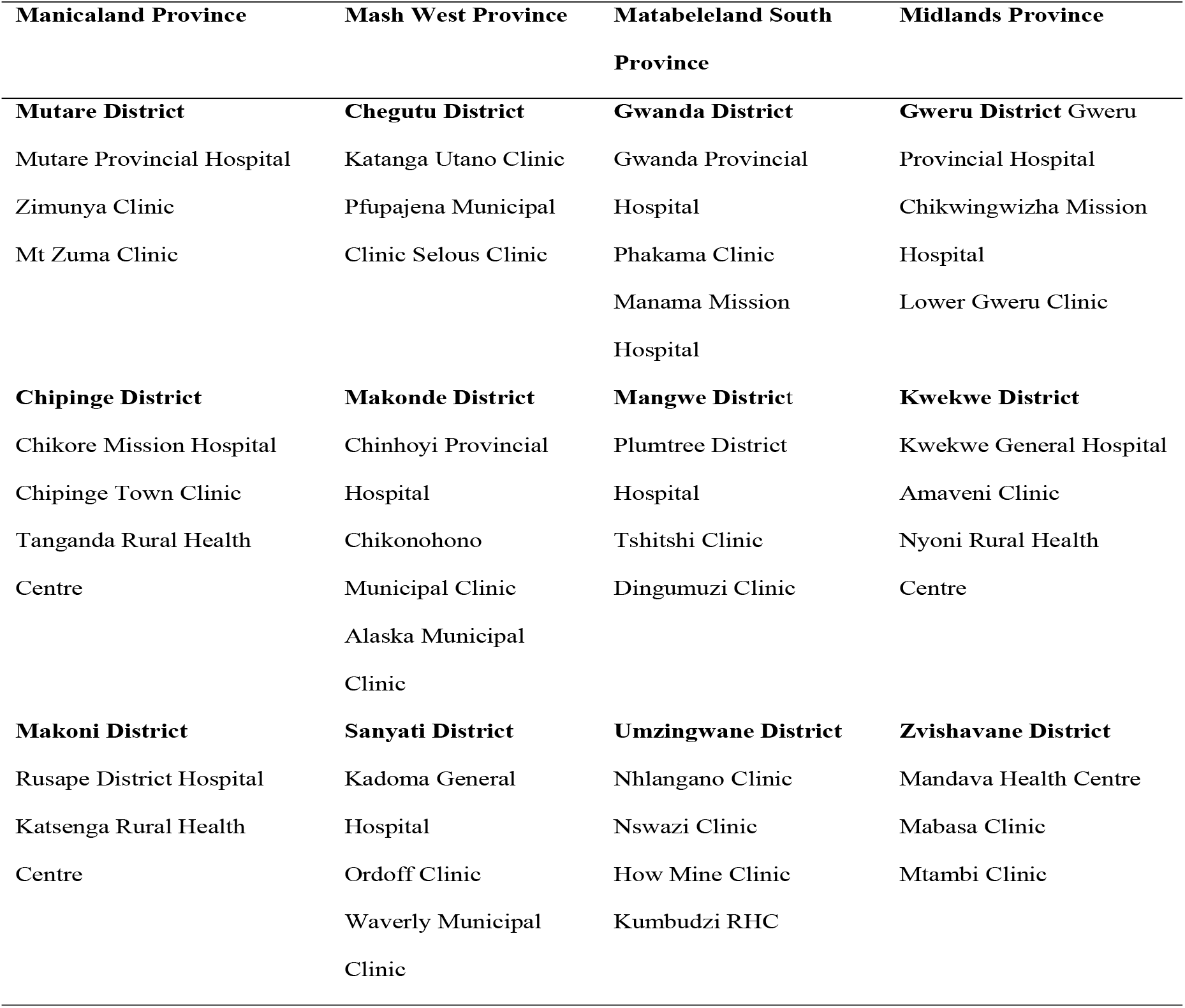
Study sites, Zimbabwe, 2024.

### Data variables, sources of data and data collection

Data were extracted from District Health Information System version 2 (DHIS2) as an excel report. The data was accessed on the 26^th^ of February 2024 for research purposes. The data was exported to EpiData Analysis version 2.2.2.186 (EpiData Association, Odense, Denmark) and Stata v14 (Stata Corporation College Station, Texas, USA) for further cleaning and analysis. The following variables were collected: HTS number, name of the facility, name of the patient, age, sex, screening for an HIV test, reason for an HIV test, HIV test result, linkage to post-test services (yes/no), and specific services linked to. No patient level data was collected.

### Analysis and statistics

Socio-demographic characteristics of participants were summarized using percentage for categorical data and mean (standard deviation) or median (interquartile range) for continuous data depending on whether they are normally distributed or not. The number and proportion with a 95% confidence interval were used to summarize all clients tested for HIV during the study period, the outcomes of the test and linkage to post-test services as documented in the respective registers.

To assess the association between risk profile, the reason for an HIV test, the result and linkage to post-test services for both negative and positive testers, an unadjusted and adjusted generalized linear model (log-binomial regression) was used. Those variables with a p-value <0.25 in the unadjusted analysis were included in adjusted models. The unadjusted and adjusted odds ratios at 5% significance levels (95%CI) were expressed as a measure of association.

### Ethics approval

Approval to conduct this study was obtained from the Ministry of Health and Childcare head office. The study was based on data collected into the DHIS2 electronic database which does not include patient identifying data. The study was therefore exempted from ethical clearances. No primary data was collected.

## Results

### Demographic characteristics

Of 23,058 patients tested for HIV, females constituted 55% (n=12,698) and 54.7% (n=12,615) patients belonged to the age group 25-49 years. In most patients, 66.3% (n=15,289) were documented to have been screened for eligibility before testing, while 33.7% were not. Among the patients tested for HIV, 63.5% (n=14,650) were retested while 1,727 tested positive, translating to a positivity ratio of 7.5%. Across the districts, Gweru recorded the highest, 15.5% (n=3,575) number of tests done whilst Mangwe recorded the least, 4.5% (n=1040) number of tests done **(Table 2)**.

**Table 2.** Clinical and demographic profile of patients, Zimbabwe, 2024. (N=23,058)

| **Variable** | **Number** | **(%)\*** |
| --- | --- | --- |
| **Total** | **23,058** | **(100)** |
| Age in years |  |  |
| ○ 15-24 | 3,524 | (15.3) |
| ○ 22-29 | 3,588 | (15.6) |
| ○ 30-34 | 5,223 | (22.7) |
| ○ 35-39 | 3,679 | (16.0) |
| ○ 40-44 | 4,315 | (18.7) |
| ○ 45-49 | 2,398 | (10.4) |
| ○ >= 50 | 331 | (1.4) |
| Gender |  |  |
| ○ Male | 10,360 | (44.9) |
| ○ Female | 12,698 | (55.1) |
| Risk Screened before testing |  |  |
| ○ Yes | 15,289 | (66.3) |
| ○ No | 7,769 | (33.7) |
| Type of HIV Test |  |  |
| ○ First Test | 7,906 | (34.3) |
| ○ Retest | 14,650 | (63.5) |
| ○ Not Documented | 502 | (2.2) |
| HIV Test Result |  |  |
| ○ Negative | 21,331 | (92.5) |
| ○ Positive | 1,727 | (7.5) |
| Testing District |  |  |
| ○ Mangwe | 1040 | (4.5) |
| ○ Sanyati | 1232 | (5.3) |
| ○ Gwanda | 1239 | (5.4) |
| ○ Chipinge | 1350 | (5.9) |
| ○ Mutare | 1254 | (5.4) |
| ○ Gweru | 3575 | (15.5) |
| ○ Kwekwe | 2443 | (10.6) |
| ○ Chegutu | 3279 | (14.2) |
| ○ Makonde | 2997 | (13.0) |
| ○ Umzingwane | 1144 | (5.0) |
| ○ Makoni | 2424 | (10.5) |
| ○ Zvishavane | 1081 | (4.7) |

\*Column percentage

### Screening, Testing Outcomes and Post-test Linkages

From a total of 23,058 patients who attended the sampled healthcare facilities, 66.3% (N=15,289) were screened for eligibility for testing. The positivity ratio among these was 8.5% (N=1,296) and almost all of them were enrolled into care (N=1,294, 99.8%). The positivity obtained among the screened patients was 75.1% of the overall positivity obtained in this study (1,296/1,727). Among the 7,769 (33.7%) clients who were not screened before testing, 431 (5.5%) tested positive and this was 24.9% of the overall positivity obtained in this study (431/1,727). Of the 7,338 (94.5%) patients who tested negative, 1,871 (25.5%) were linked to HIV prevention services **(Figure 2)**.

**Figure 2.**
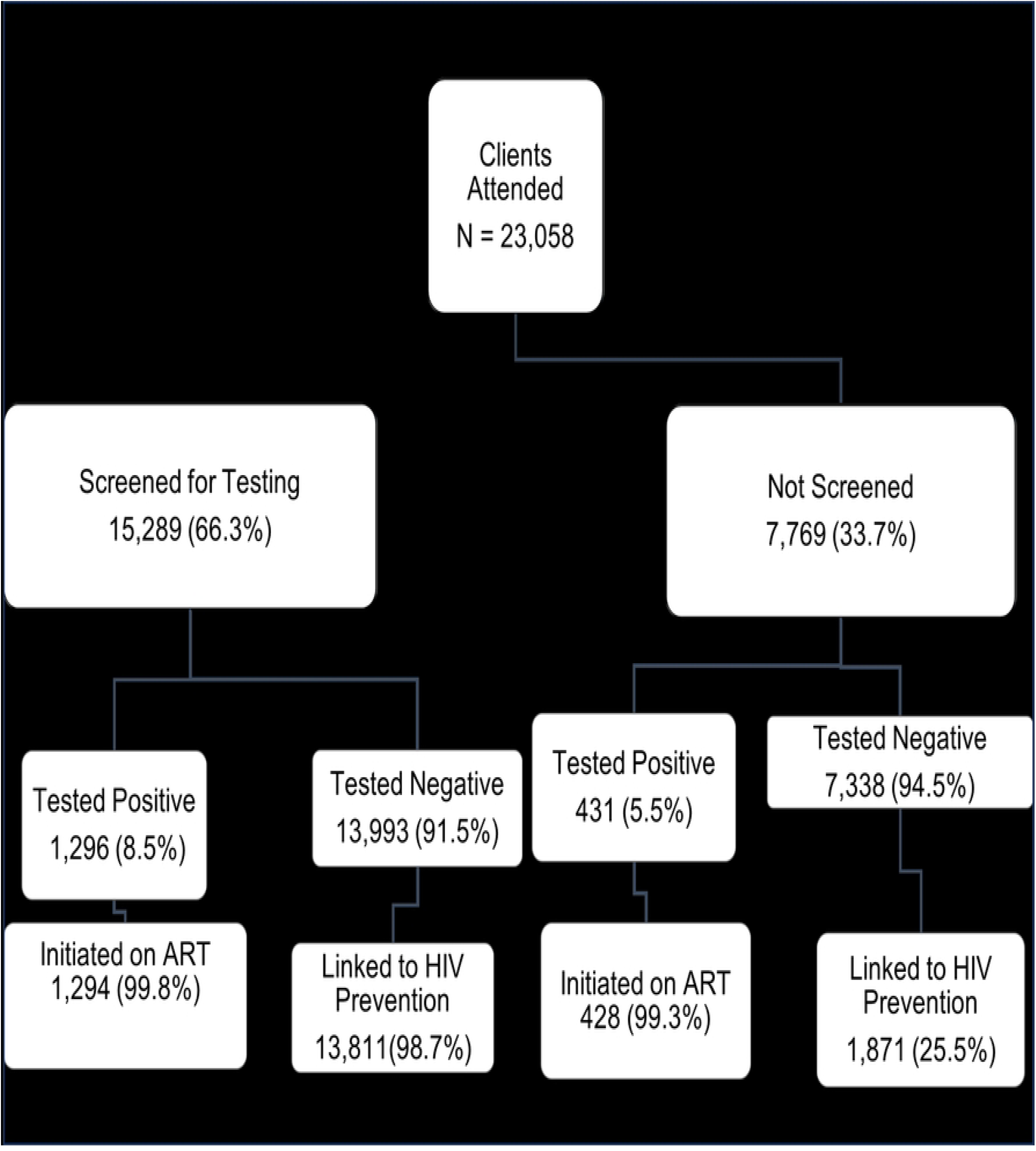
HIV Testing and Post-test Linkages, Zimbabwe, 2024.

### HIV Positivity and Linkage

The overall positivity ratio obtained in this study was 7.5% (1,727/23,058). The 45–49-year category was 3.6 times more likely to test positive (a95%CI:2.67,4.90). Males were 3.09 times more likely to test HIV positive in adjusted analysis (a95%CI: 2.74, 3.49), from an 8% (n=912) positivity ratio. The first tests were 65% more likely to test HIV positive (a95%CI: 1.43, 1.91) whilst patients who were screened before testing were 3.89 times more likely to link to at least 1 HIV prevention service (a95%CI: 3.05, 4.97). **(Table 3.)**

**Table 3.**
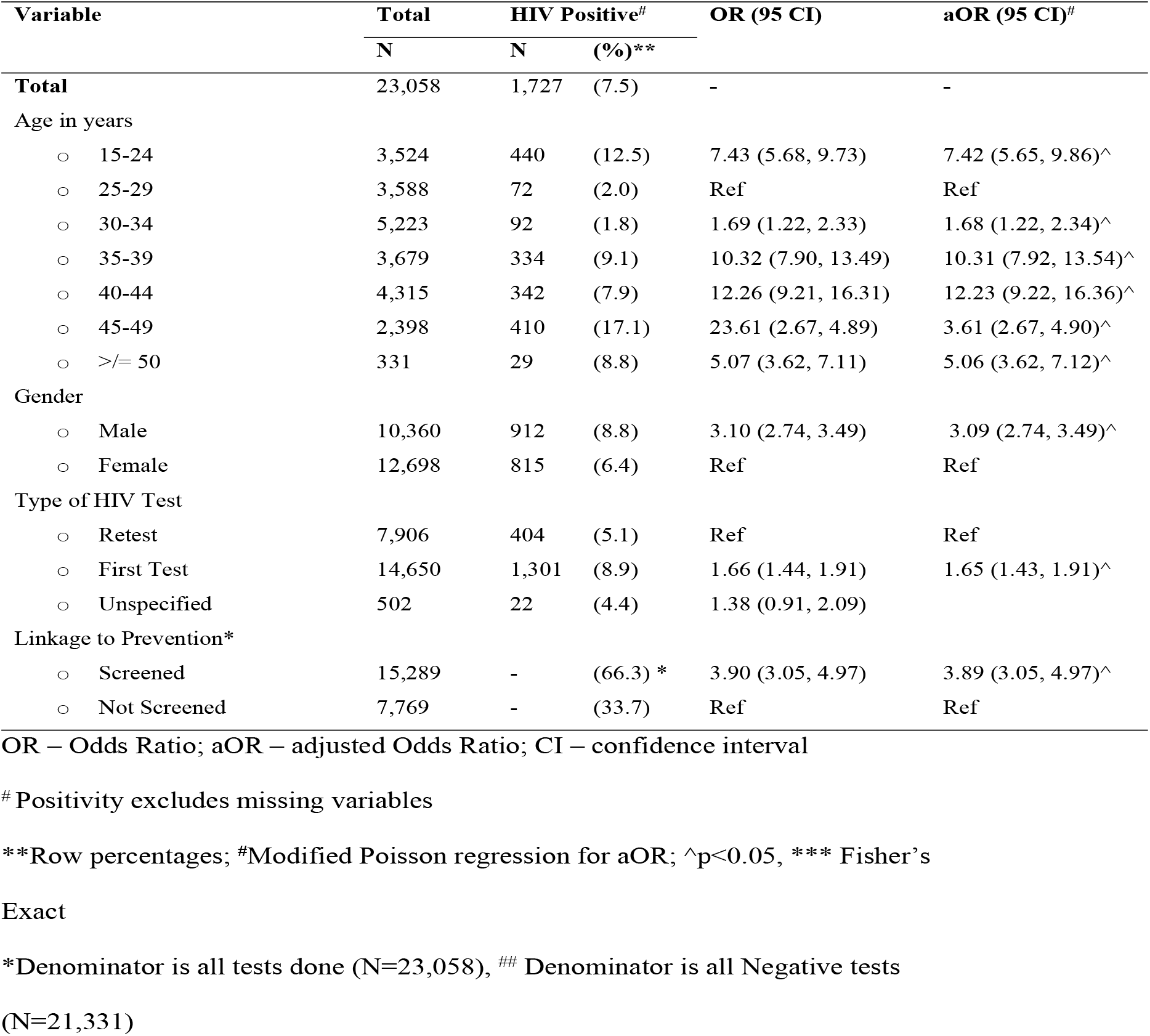
Factors associated with HIV Positivity among patients who tested for HIV, Zimbabwe, 2024. (N=23,058)

## Discussion

A key finding in this study is that the status-neutral approach to HIV testing complements targeted HIV testing, enabling prioritized testing and linkage to HIV prevention and treatment services.

### Strengths

The availability of primary source documents, such as HTS registers at all visited facilities facilitated the data abstraction process. In addition, the sampled 36 health facilities provided a large sample size that enabled us to draw inferences on the population of the country.

### Limitations

Discrepancies between data abstracted from HTS registers, monthly summaries and DHIS2 during data triangulation exposed data entry or computation errors that could be rectified by onsite data analysis, and cascade generation.

### Interpretation of key findings

This study provided important insights into the role of status-neutral testing in targeted testing in Zimbabwe.

First, patients who were screened for their risk for HIV infection before testing scored a high positivity ratio of 8.5% compared with those who were not screened (5.5%). The risk screening was done using a standardized screening algorithm which is part of standard service delivery by the country, in determining eligibility for an HIV test [17].

Further, the screened patients contributed as high as 75% of the overall positivity obtained, when the calculation combined with those not screened. This indicates the relevance of targeted testing in Zimbabwe, in the context of a generalized epidemic being managed against a backdrop of declining funding for HIV, characterized by sporadic stockouts of testing commodities for HIV testing. This finding is consistent with previous studies that underscore the importance of an algorithm to determine risk and prioritize clients for HIV testing, whilst offering self-screen to those at low risk [17, 22–24] To remain on course of achieving and sustaining the targets for case identification, it is therefore imperative to effectively implement an algorithm that aids health workers in prioritizing patients who are most likely to test HIV positive

Secondly, 98.7% (n=13,811) of the clients who tested HIV negative following risk screening were linked to HIV prevention services. In multivariate analysis, the probability of linkage among HIV-negative screened clients was 3.89 (a95%CI:3.05, 4.97). This finding indicates that screening clients for testing assists in focussed HIV prevention linkage among the clients who test negative. Our findings are consistent with the tenets of the status-neutral concept of equal emphasis on linkage to prevention and treatment [6, 7, 25, 26] but vary with the placing of risk assessment where other studies place it after the test, rather than before the test. Placing the risk screening stage after the test results in an increased number of clients in the post-test stage with an unknown risk profile requiring risk screening. The increased numbers, particularly in our context of high numbers and high-frequency testing may affect the vigilance with which the screening process is done [27, 28].

In this study, we found the utility of risk screening being done before conducting the test for the status-neutral concept to effectively complement the targeted testing framework which reduces the high frequency of testing which does not correspond with the risk profile. Implementing the status-neutral concept without embedding it into targeted testing departs from the standardized retesting algorithm which restricts the highest frequency of HIV testing to once every three months, for people at ongoing risk for HIV infection [7, 29].

Third, 63.5% of the clients who tested for HIV were retests (n=14,650), and yet first tests were 65% more likely to test HIV positive (a95%CI: 1.43, 1.91), adjusted for age and sex. Most tests being retested may be suggestive of “over-testing” or high-risk perception, particularly given the low positivity ratios obtained [4]. Clients who test HIV negative at contact are retested annually if they fall into the general population category and retested 3 monthly if they are at ongoing risk for HIV transmissions, such as sero-different couples and those on PrEP, according to the national retesting algorithm [30].

Finally, men were 3.09 times more likely to test HIV positive (a95%CI:2.74, 3.49) in adjusted analysis despite contributing 44.9% (10,360/23,058) of the tested population. This finding implies that fewer men test, from which most test positive. This finding corroborates previous studies and information in the public domain [31, 32]. Further, men are also less likely to adhere to treatment and more likely to have unfavourable treatment outcomes, with attributable factors that include gender norms [30, 33].

### Implications for policy and practice

Targeted testing is the mainstay of HTS programming to achieve epidemic control. Complemented with status neutral approach to HIV testing will bring out a double-edged sword, one that prioritizes testing and linkage to both prevention and treatment. The concept needs to be vigilantly implemented to expedite epidemic control, whilst preventing new infections through linkage to prevention as well.

## Conclusions

Targeted testing and status-neutral testing are related concepts which are complimentary. Simultaneously applied, the concepts facilitate active identification of people living with HIV to meet case identification targets, through prioritizing high-risk individuals for testing, followed by arresting ongoing transmission of HIV through effective linkage to HIV prevention and treatment. This approach will facilitate the best usage of limited resources, particularly in low to medium countries.

## Data Availability

All relevant data are within the manuscript and its Supporting Information files.

## Acknowledgements

I acknowledge several individuals and institutions that made this study a success. Special gratitude goes to my academic supervisors, Professor M. Tshimanga, Dr J. Chirenda and Dr K. Takarinda, The Director of AIDS & TB Unit, Dr O. Mugurungi and the entire HTS team for their support and prodding during this study.

